# Genome-wide association study reveals shared and distinct genetic architecture underlying fatty acid and bioactive oxylipin metabolites in the Hispanic Community Health Study/Study of Latinos (HCHS/SOL)

**DOI:** 10.1101/2024.05.21.24307719

**Authors:** Carolina G. Downie, Heather M. Highland, Mona Alotaibi, Barrett M. Welch, Annie Green Howard, Susan Cheng, Nick Miller, Mohit Jain, Robert C. Kaplan, Adam G. Lilly, Tao Long, Tamar Sofer, Bharat Thyagarajan, Bing Yu, Kari E. North, Christy L. Avery

**Affiliations:** Department of Epidemiology, University of North Carolina at Chapel Hill, Chapel Hill, NC; Division of Pulmonary, Critical Care and Sleep Medicine, University of California, San Diego, San Diego, CA; School of Public Health, University of Nevada, Reno, Reno, NV; Department of Biostatistics, University of North Carolina at Chapel Hill, Chapel Hill, NC; Department of Cardiology, Smidt Heart Institute, Cedars-Sinai Medical Center, Los Angeles, CA; Sapient Bioanalytics, San Diego, CA; Departments of Medicine and Pharmacology, University of California, San Diego, San Diego, CA; Department of Epidemiology and Population Health, Albert Einstein College of Medicine, Bronx, NY; Public Health Sciences Division, Fred Hutchison Cancer Center, Seattle, WA; Department of Sociology, University of North Carolina at Chapel Hill, Chapel Hill, NC; CardioVascular Institute (CVI), Beth Israel Deaconess Medical Center, Boston, MA; Department of Biostatistics, Harvard T.H. Chan School of Public Health, Boston, MA; Department of Laboratory Medicine and Pathology, University of Minnesota Medical Center, Minneapolis, MN; Department of Epidemiology, Human Genetics, and Environmental Sciences, The University of Texas Health Science Center at Houston School of Public Health, Houston, TX

## Abstract

Bioactive fatty acid-derived oxylipin molecules play key roles in mediating inflammation and oxidative stress, which underlie many chronic diseases. Circulating levels of fatty acids and oxylipins are influenced by both environmental and genetic factors; characterizing the genetic architecture of bioactive lipids could yield new insights into underlying biological pathways. Thus, we performed a genome wide association study (GWAS) of n=81 fatty acids and oxylipins in n=11,584 Hispanic Community Health Study/Study of Latinos (HCHS/SOL) participants with genetic and lipidomic data measured at study baseline (58.6% female, mean age = 46.1 years, standard deviation = 13.8 years). Additionally, given the effects of central obesity on inflammation, we examined interactions with waist circumference using two-degree-of-freedom joint tests. Heritability estimates ranged from 0% to 47.9%, and 48 of the 81oxylipins and fatty acids were significantly heritable. Moreover, 40 (49.4%) of the 81 oxylipins and fatty acids had at least one genome-wide significant (*p*< 6.94E-11) variant resulting in 19 independent genetic loci involved in fatty acid and oxylipin synthesis, as well as downstream pathways. Four loci (lead variant minor allele frequency [MAF] range: 0.08-0.50), including the desaturase-encoding *FADS* and the OATP1B1 transporter protein-encoding *SLCO1B1*, exhibited associations with four or more fatty acids and oxylipins. The majority of the 15 remaining loci (87.5%) (lead variant MAF range = 0.03-0.45, mean = 0.23) were only associated with one oxylipin or fatty acid, demonstrating evidence of distinct genetic effects. Finally, while most loci identified in two-degree-of-freedom tests were previously identified in our main effects analyses, we also identified an additional rare variant (MAF = 0.002) near *CARS2*, a locus previously implicated in inflammation. Our analyses revealed shared and distinct genetic architecture underlying fatty acids and oxylipins, providing insights into genetic factors and motivating future multi-omics work to characterize these compounds and elucidate their roles in disease pathways.

## Introduction

Chronic inflammation is a pathological feature underlying many chronic diseases,^1–5^ and inflammatory responses involve a variety of so-called “inflammatory mediators” that include both signaling proteins (e.g., cytokines, chemokines) and bioactive lipids such as oxylipins.^6^ Oxylipins are derived from the oxygenation of precursor mono- and polyunsaturated fatty acids through cascading pathways mediated by cytochrome P450 (CYP), lipoxygenase (LOX), and cyclooxygenase (COX) enzymes, or through other enzymatic and non-enzymatic reactions^7^ (**Supplementary** Figure 1), and play a variety of roles mediating inflammation and oxidative stress.^8,9^ Given the role of inflammation in many chronic diseases, there is growing interest in oxylipin-targeted therapies.^10^ Oxylipins and their precursor fatty acids are influenced by both environmental and genetic factors.^11–13^ Thus, characterizing the genetic architecture of oxylipins and related fatty acids could yield new insights into underlying biological pathways that could inform drug targets.^11^

Genetic analyses of several polyunsaturated fatty acids from which many oxylipins are derived have identified multiple variants at loci encoding desaturase and elongase enzymes (e.g., *FADS, ELOVL2*).^14–18^ Moreover, previous associations between the LOX-encoding *ALOX12* locus and 12-HETE oxylipins have been reported, consistent with the role that LOX enzymes play in the derivation of these metabolites.^12,19^ Still, few analyses of the genetic architecture of oxylipins have been conducted on a large scale. This is due in part to historical challenges in quantifying and annotating oxylipin species: hundreds of oxylipins with distinct structures and stereochemistry can be derived from fatty acids.^8^ However, recent advancements in mass spectrometry-based metabolomics and annotation approaches have enabled the improved detection, quantification, and annotation of oxylipins.^20–22^

To address this research gap, we estimated the heritability and performed a genome-wide association study (GWAS) of n=81 oxylipins and related fatty acids quantified via directed, non-targeted liquid chromatography-mass spectrometry (LC-MS) in the Hispanic Community Health Study/Study of Latinos (HCHS/SOL). Moreover, motivated by the fact that central adiposity is associated with other markers of low-grade inflammation (e.g., C-reactive protein and interleukin-6)^23–26^ and may influence oxylipin biosynthesis,^27^ we also extended our models to include interactions with waist circumference and performed two-degree-of-freedom joint tests, increasing our power to identify additional loci that have both main and interaction effects.^28,29^ Our findings reveal both shared and distinct genetic architecture of modest-to-moderately heritable circulating fatty acids and oxylipins, providing insights into the biological pathways underlying these metabolites.

## Methods

### Hispanic Community Health Study/Study of Latinos (HCHS/SOL)

HCHS/SOL is a multi-center, community-based cohort study of Hispanic/Latino adults in the United States designed to study the incidence and prevalence of, and risk factors for, multiple chronic diseases, including cardiovascular diseases, kidney, lung and liver diseases.^30^ A total of 16,415 participants of Mexican, Puerto Rican, Dominican, Cuban, and Central and South American background aged 18-74 years were recruited from four U.S. cities between 2008 and 2011: Bronx, NY; Chicago, IL; Miami, FL; and San Diego, CA. Households were selected via two-stage sampling within census block groups.^31^ At study baseline (2008-2011), participants received standardized examinations and interviewer-administered questionnaires, with follow-up examinations in 2015-2017 and 2020-2023. Informed consent was obtained from all participants, and the study was conducted in accordance with the principles of the Declaration of Helsinki. Approval from the University of North Carolina at Chapel Hill Institutional Review Board was obtained for this work.

### Oxylipin profiling, annotation, and quality control

Plasma oxylipin profiling was conducted by Sapient Bioanalytics, using a rapid liquid chromatography-mass spectrometry (rLC-MS) methodology that has been described in detail previously^22^ (**Supplementary Methods**). Oxylipins and fatty acids were annotated by aligning experimental accurate mass-to-charge ratio (m/z), retention time (RT) and MS2 spectra with an in-house curated library of commercial standards as well as alignment with previously reported structural isomers.^1^ These annotations often indicated multiple potential chemically related compound mappings, due to compounds with similar chemical formulas and mass-to-charge ratios co-eluting at the same time. Furthermore, oxylipin metabolites with chemical structures that aligned with previously reported structural isomers for which biological pathways were unknown were annotated as “putative known oxylipins.” Likewise, oxylipins with chemical structures that did not align to any previously reported standards or structural isomers and for which biological pathways were unknown were annotated as “putative novel oxylipins.”

In this study, we restricted our analysis to n=81 metabolites annotated as oxylipins and fatty acids based on the procedure described above. To facilitate interpretation of analyses, for the oxylipin metabolites, we defined precursor fatty acid groupings based on literature searches describing the biosynthetic pathways of oxylipin compounds as well as online resources such as PubChem (https://pubchem.ncbi.nlm.nih.gov), the Human Metabolome Database (https://hmdb.ca), and Cayman Chemical (https://www.caymanchem.com). When annotations indicated multiple potential compound mappings due to compounds with similar chemical formulas and mass-to-charge ratios co-eluting at the same time, we grouped them based on any of the precursor fatty acids from which they may have been derived (e.g., “arachidonic acid or eicosapentaenoic acid”). Moreover, fatty acids were grouped based on the class of fatty acids to which they belonged: polyunsaturated fatty acids, monounsaturated fatty acids, or saturated fatty acids.

The percent missingness across all 81 metabolites ranged from 0% −23.4%, with the majority of metabolites (93.8%) missing less than 2% of all observations. Metabolite observations with missing data were imputed from a random uniform distribution between the minimum measured metabolite value/10 and the minimum measured metabolite values. All metabolite values were subsequently log_2_-transformed to reduce the impact of extreme outlier values.^32^

### Genotyping and quality control

HCHS/SOL participants were genotyped on the Infinium Expanded Multi-Ethnic Genotyping Array (MEGA)^33^, and imputed to the TOPMed R2 (Freeze 8) imputation panel using IMPUTE v2.3.2 (**Supplementary Methods**). Variants with an imputation quality score < 0.4 or effective sample size < 20, calculated as 2×MAF×(1-MAF)×N×Q, where MAF is minor allele frequency, Q is imputation quality, and N is sample size, were excluded from main effect analyses. For joint effect analyses, an effective sample size filter of 30 was applied, where the N used in effective sample size calculations was the sex-stratified sample size (N_females_ = 6725, N_males_ = 4753). All genomic coordinates are reported on GRCh38.

### Spearman correlations

Because Spearman correlations are more robust to outliers and non-normal distributions than Pearson correlations,^34^ we computed Spearman correlations of the n=81 oxylipins and fatty acids to determine their correlation structure.

### Heritability analysis

We estimated SNP-based heritability, or the proportion of variance explained for each metabolite due to the additive effect of genotyped common variants, and corresponding 95% confidence intervals, using Haseman-Elston regression, as previously described for the HCHS/SOL study.^35^ For this analysis, we used a kinship matrix calculated using autosomal variants genotyped on an Illumina custom array (SOL HCHS Custom 15041502 B3), consisting of the Illumina Omni 2.5M array (HumanOmni2.5-8v1-1) and approximately 150,000 custom variants,^36^ and then imputed to the 1000 Genomes Phase 3 reference panel.

### Main effects GWAS

The log_2_-transformed metabolite values were regressed on age, age^2^, sex, study recruitment site, and self-identified background, and residuals from these models were subsequently inverse rank normalized to yield asymptotically normal marginal distributions.^37^ Genetic association analyses of the inverse rank normalized residuals were conducted using generalized estimating equations via SUGEN,^38^ adjusting for 10 ancestral principal components, which were calculated using Plink.

To account for multiple testing of 81 correlated oxylipins and fatty acids, we used a modified Bonferroni correction based on the effective number of independent tests.^39^ Briefly, we computed the number of principal components necessary to explain 99.5% of metabolite variability (N_effective_= 72), yielding a multiple testing-corrected p-value threshold of 5E-09/72 = 6.94E-11. We used EasyStrata software^40^ to identify independent loci and corresponding top variants, using 500 KB as the physical position threshold for clumping. We used the Open Targets Genetics tool (https://genetics.opentargets.org) to annotate the nearest genetic locus for each of our top variants.

### Main effects conditional analyses

We performed stepwise exact conditional analyses to identify additional independent secondary signals for each genome-wide significant locus identified in the main effects GWAS. Briefly, we included the top genome-wide significant variant at each significant locus as a covariate in our SUGEN analysis and iterated until no novel significant secondary signals (*p* < 6.94E-10) remained. We then estimated the proportion of variance in the phenotype explained (PVE) by a given variant for each independent index and secondary variants using the following equation, as previously described^41,42^:

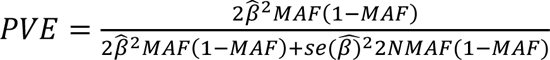

### Joint effects GWAS

To increase the power to identify additional loci in which genetic variants exhibit both a main effect and an interaction effect,^28,29^ we also conducted joint two-degree-of-freedom tests accounting for waist circumference as a measure of central adiposity. Because sex-specific waist circumference thresholds have been recommended for defining central obesity,^43,44^ we conducted sex-stratified joint effect analyses, including inverse rank normalized residuals of waist circumference adjusted for body mass index (BMI) as an interaction term in our analyses. Waist circumference, weight, and height were measured using standardized protocols at the baseline study visit. Waist circumference adjusted for BMI was calculated in sex-stratified analyses, regressing waist circumference on age, age^2^, and BMI, and then inverse rank normalizing the residuals.^45^ We then meta-analyzed the p-values for the joint variant and variant x waist circumference effect (“joint p-value”) using METAL,^46^ weighting by sample size. All figures were generated using R version 4.1.0, using the colorblind-friendly color palette from the following resource: https://zenodo.org/record/3381072.

## Results

There were n=11,610 SOL participants with measured genetic and metabolite data at the baseline visit. After excluding n=26 participants with missing self-reported background group, the final main effects analysis sample was n=11,584. This analytic sample was 58.6% female, and the mean age of participants was 46.1 years (standard deviation [SD]=13.8). After excluding an additional n=106 with missing BMI-adjusted-waist circumference, the final joint effect analysis sample was n_Females_=6,725 and n_Males_=4,753 (total n=11,478). The mean waist circumference for females was 97.6 centimeters (SD=14.2 cm), and the mean waist circumference for males was 99.0 centimeters (SD=13.3 cm).

### Spearman correlation of fatty acids and oxylipins

Spearman correlations of the log_2_-transformed values of the 81 oxylipins and fatty acids ranged between −0.27 to 0.98 (mean=0.21), with the strongest correlations observed between fatty acids (range: −0.22 to 0.90, mean=0.34) (**Figure 1, Supplementary** Figure 2). Among the oxylipins, correlations varied both within and between precursor fatty acid groupings and were highest for oxylipins derived from linoleic acid (range: −0.12 to 0.70, mean=0.28) and lowest for oxylipins putatively derived from arachidonic acid or eicosapentaenoic acid (range: −0.06 to 0.18, mean=0.07) (**Figure 1, Supplementary** Figure 2).

**Figure 1.**
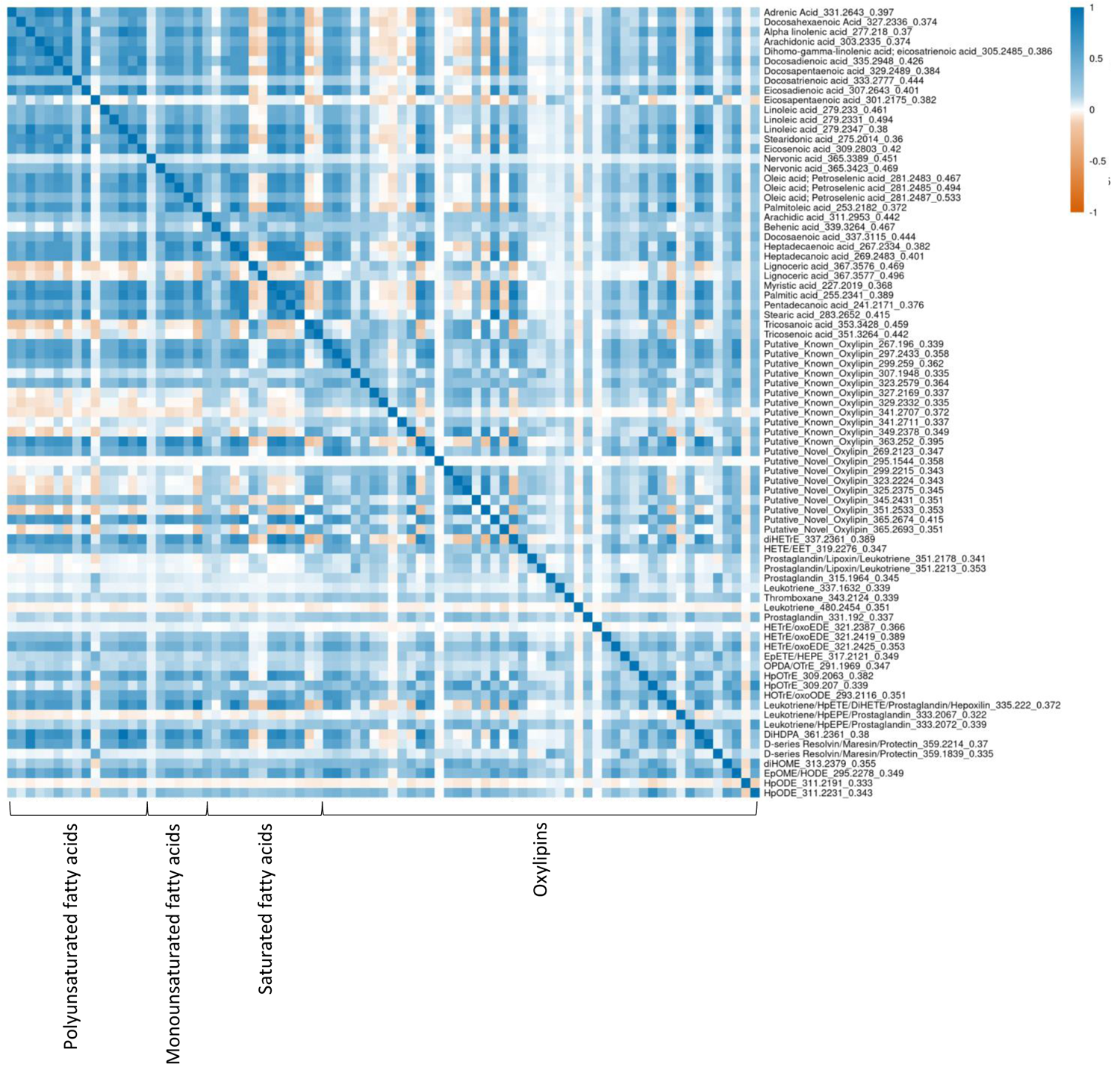
Spearman correlation matrix of log2-transformed values of n=81 putative fatty acids and oxylipin metabolites. Positive correlations are displayed in blue, negative correlations are displayed in orange. Metabolites are annotated with the putative general class of compound to which they belong, their measured mass-to-charge ratio (m/z), and their retention time (RT) in the form: metabolite name_m/z_RT. When annotations indicated multiple potential compound mappings due to compounds with similar chemical formulas and mass-to-charge ratios co-eluting at the same time, we have listed all of them, separated by backslashes (e.g., HETE/EET). Metabolites are ordered in the matrix based on these general groupings, with putative free fatty acids first (polyunsaturated fatty acids, monounsaturated fatty acids, saturated fatty acids), followed by oxylipin metabolites.

### Heritability of fatty acids and oxylipins

Forty-eight of the 81 (59.3%) fatty acids and oxylipins exhibited significant (*p* < 0.05) heritability (significant h_SNP_ ^2^ range: 4.6% to 47.9%), the magnitude of which varied across the 34 fatty acids and 47 oxylipins (**Supplementary Table 1**). Among metabolites with significant heritability, fatty acids generally exhibited higher heritability (mean h_SNP_^2^ = 12.6%, SD = 10.2%, range: 5.0% to 47.9%) than oxylipins (mean h_SNP_^2^=8.2%, SD=3.4%, range: 4.6% to 16.9%) (**Figure 2, Supplementary Table 1**). Among oxylipins, heritability estimates were generally consistent across fatty acid precursor groups.

**Figure 2.**
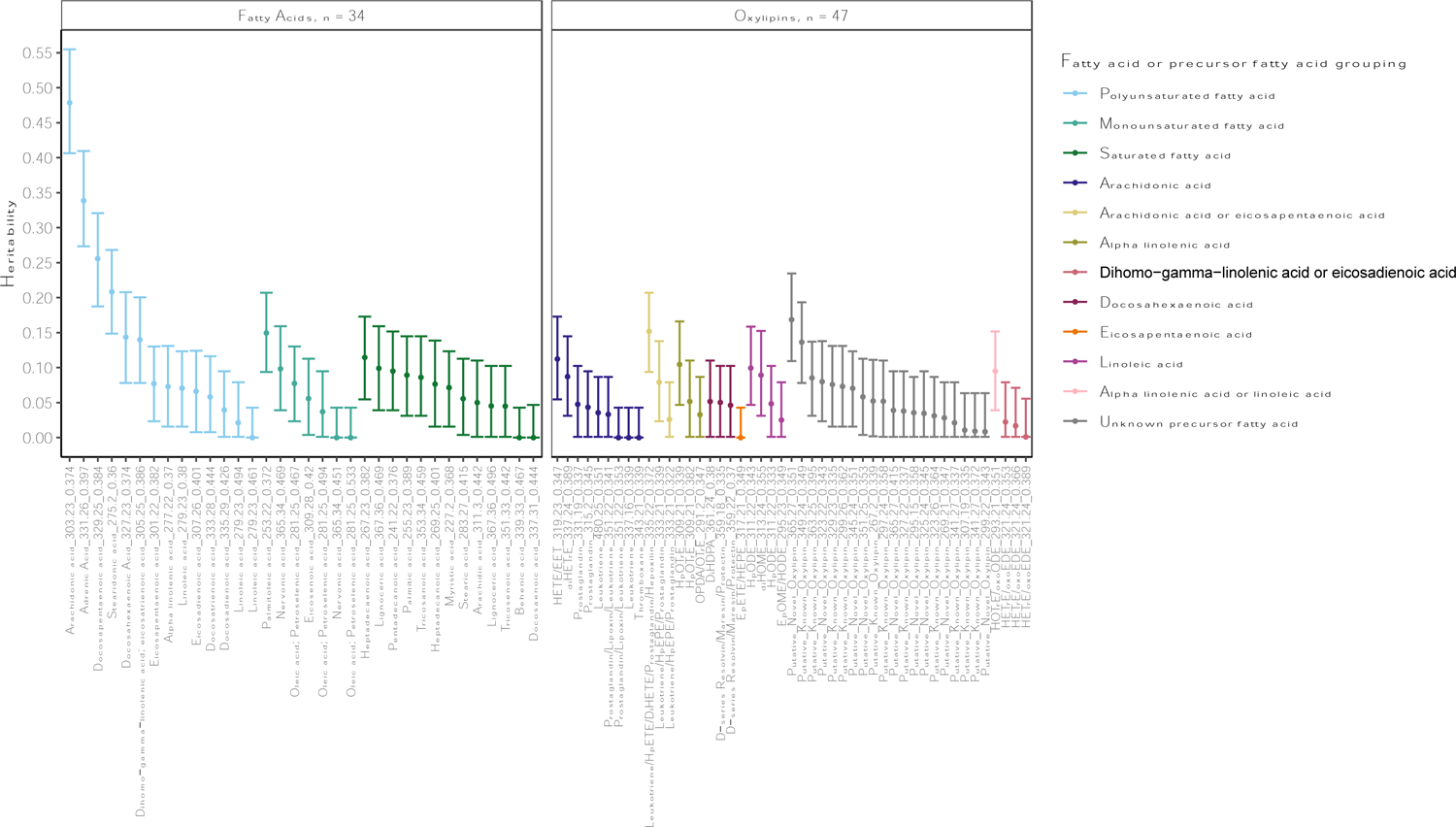
Estimated proportion of variance due to kinship (heritability) and 95% confidence intervals of n=81 putative fatty acids and oxylipin metabolites. Metabolites are annotated with the putative general class of compound to which they belong, their measured mass-to-charge ratio (m/z), and their retention time (RT) in the form: metabolite name_m/z_RT. Estimates are colored by the putative grouping to which the metabolite belongs: polyunsaturated fatty acid, monounsaturated fatty acid, saturated fatty acid, or the putative fatty precursor from which the putative oxylipin is derived. Putative novel and known oxylipins are grouped in “Unknown precursor fatty acid” grouping.

### Main effects GWAS

Of the 81 fatty acids and oxylipins, 40 (49.4%) had at least one genome-wide significant (*p* < 6.94E-11) variant. These metabolites included 33 of the 48 (68.8%) significantly heritable fatty acids and oxylipins (**Supplementary Table 1**). After accounting for variants that mapped to the same locus, there were 19 independent loci, and 55 unique metabolite-locus pairs (**Figure 3a, Supplementary Table 2, Supplementary** Figure 3). The top variants were all common (minor allele frequency (MAF) > 0.01), and all of these variants or their high linkage disequilibrium (LD) proxies have been previously associated with metabolites, though not necessarily oxylipins (**Supplementary Table 2**). As previously reported, allele frequencies for several of the loci we identified varied across reference continental ancestry populations, including *FADS*^18^, the *CYP* supergene family,^47^ *SLCO1B1,*^48,49^ *FUT2,*^50^ and *MCM6.*^51^ Moreover, *ADH1A* top variant rs28600890 was monomorphic in 1000 Genomes European and South and East Asian reference populations (**Supplementary Table 2, Supplementary** Figure 4).

**Figure 3.**
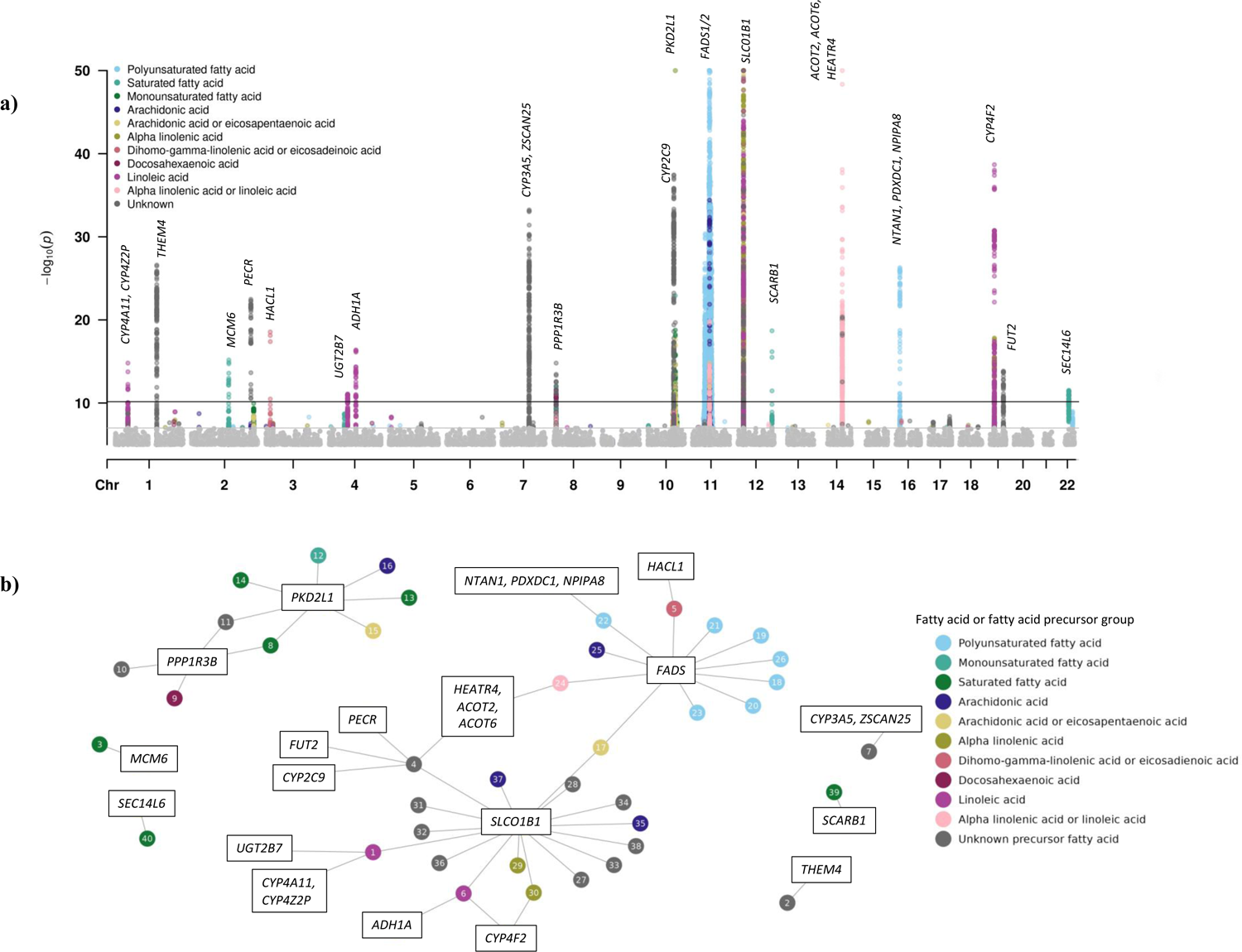
Summary of unique loci identified in main effects GWAS. **(a)** Manhattan plot of unique loci identified in main effects GWAS, color coded by the putative grouping to which the metabolite belongs: polyunsaturated fatty acid, monounsaturated fatty acid, saturated fatty acid, or the putative fatty precursor from which the putative oxylipin is derived. Oxylipins annotated as ‘putative novel’ or ‘putative known’ are colored in gray, and precursor fatty acid group is marked as “Unknown precursor fatty acid.” -log10 p-values were capped at 50 to aid visualization of results (maximum -log10pvalue observed was 262, for arachidonic acid associated with *FADS2*). Some annotations included multiple putative compounds which are derived from different fatty acid precursors, so both are listed (e.g., dihomo-gamma-linolenic acid or eicosadienoic acid; arachidonic acid or eicosapentaenoic acid). Black line indicates genome-wide significance threshold (5E-09/72 = 6.94E-11); all variants with p-value >1E-7 are colored in gray to improve visualization of results. **(b)** Network plot of the 56 unique metabolite-locus associations to illustrate pleiotropic and distinct genetic effects. Genetic loci are in boxed labels, metabolites are colored based on the putative fatty acid or precursor fatty acid grouping from which the oxylipin is derived. Metabolites are numbered (1-40) and these numbers correspond to the “metabolite id number” column in Supplementary Table 2.

### Shared Genetic Architecture

The genetic architecture of the n= 40 fatty acids and oxylipins with at least one genome-wide significant variant was characterized by both shared and distinct genetic effects. Of the 20 identified loci, four pleiotropic loci (*SLCO1B1, FADS, PKD2L1,* and *PPP1R3B*) were associated with four or more metabolites (**Figure 3b**). These loci accounted for 38 (69.1%) of the 55 unique metabolite-locus associations, included common variants (MAF range: 0.08 – 0.50, mean: 0.33), and generally belonged to similar fatty acid or precursor fatty acid groupings, though *PKD2L1* was associated with saturated and monounsaturated fatty acids as well as oxylipins (see **Figure 3b, Supplemental Table 5**). Moreover, most of the oxylipins and fatty acids (71.4%) associated with these pleiotropic loci did not have significant associations with any other additional loci.

Furthermore, there were 12 highly correlated metabolite pairs (Spearman correlation coefficient > 0.80) for which one metabolite had a genome-wide significant association with one of these pleiotropic loci, but the other did not have a genome-wide or near genome-wide significant association for the same locus (see **Supplementary Table 3**). For example, a fatty acid annotated as dihomo-gamma-linolenic acid/eicosatrienoic acid (m/z = 305.25, RT=0.386) was associated with both the *FADS* and *NTAN1* loci (*p_FADS_*= 1.20E-22, *p_NTAN_ =* 5.03E-27), but correlated eicosadienoic acid (Spearman correlation = 0.81, m/z=307.26, RT=0.401) was associated with neither locus (*p_FADS_* = 0.71, *p_NTAN_ =* 0.57). This may reflect non-genetic drivers, such as diet, of the correlations between these fatty acids and oxylipins.

These four pleiotropic loci also showed little evidence of allelic heterogeneity (**Supplementary Tables 2, 4**). For example, lead missense variant rs4149056 was shared by all 16 *SLCO1B1-*associated metabolites, and variant rs11045856 was identified as the top secondary variant in all conditional analyses. Similarly, lead variant rs174570 was shared by five of the 11 *FADS*-associated metabolites, including three of the four oxylipins associated with *FADS*. Variants underlying a shared genetic architecture also explained the highest observed estimated percentage of trait variance. For example, *FADS* variant rs174564 explained 9.4% of arachidonic acid (m/z = 303.23, RT=0.374) trait variance and *SLCO1B1* variant rs4149056 explained 6.78% of HpODE (m/z =311.22, RT=0.343) trait variance (**Supplementary Table 2**).

### Distinct Genetic Architecture

The remaining 15 significant loci were each associated with three or fewer metabolites, including 13 only associated with a single metabolite (*MCM6; SCARB1; SEC14L6*; *THEM4*; *CYP3A5, ZSCAN25; ADH1A; FUT2; PECR; CYP2C9; HACL1; NTAN1, PDXDC2, NPIPA8; UGT2B7; CYP4A11, CYP4Z2P*) (**Figure 3b**, **Supplementary Table 2**). Some of these loci belonged to shared gene families, such as the four CYP loci associated with only two (*CYP4F2*) or one (*CYP2C9; CYP4A11, CYP4Z2P; CYP3A5, ZSCAN25*) metabolites. Lead variants at these loci were common (MAF range: 0.18-0.25, mean = 0.23) and demonstrated evidence of distinct genetic effects within a broad biosynthetic pathway among modestly correlated metabolites (Spearman correlation range: 0.11 - 0.74) (**Supplementary Table 5**).

We also identified unique genetic loci for metabolites that were highly correlated. For example, pentadecanoic acid (m/z = 241.22, RT=0.376) was highly correlated with myristic acid (m/z = 227.20, RT=0.368) (Spearman correlation = 0.83), but only pentadecanoic acid was significantly associated with the *MCM6* locus (rs4988235, MAF = 0.21, *p_pentadecanoic acid_* = 6.57E-16, *p_myristic acid_*= 3.91E-04) (**Supplementary Table 3**). For 13 of the 15 loci associated with only one or two metabolites, the lead variant (MAF range: 0.02 – 0.45, mean: 0.22) explained less than 1% of the trait variance (**Supplementary Table 2**).

### Unique putative novel oxylipin was associated with 5 genetic loci

Thirty-seven of the 40 (92.5%) fatty acids and oxylipins were only significantly associated with one or two genetic loci. However, one oxylipin annotated as a “putative novel oxylipin” (m/z = 365.27, RT=0.351), exhibited the highest heritability of all oxylipins (h_SNP_ ^2^ = 16.9% [95% CI: 10.9% - 23.5%]) and was associated with five loci, including shared genetic locus *SLCO1B1*. Three of these loci were specific only to this metabolite: *PECR* (missense variant rs9288513, MAF = 0.13), *CYP2C9* (rs9332172, MAF = 0.17), and *FUT2* (stop gained variant rs601338, MAF = 0.36). Together, these five lead variants explained 5.0% of trait variance. (**Supplementary Table 2**).

### Joint effects GWAS

Joint two-degree-of-freedom tests, which were conducted to increase the power to identify additional loci in which genetic variants exhibit both a main effect and interaction effect with waist circumference,^28,29^ identified 35 fatty acids and oxylipins with at least one genome-wide significant locus (*p_joint_* < 6.94E-11) (**Supplementary Table 6**). The chromosome 13 *CARS2* locus, which was associated with a leukotriene (m/z=480.25, RT=0.351), was the only locus not previously identified in our main effect GWAS (*p_main effects_* = 0.16). The *CARS2* top variant rs183091953 was significant among females (joint effects *p_joint, females_* = 2.00E-15), but it did not pass the effective N > 30 filter in males (**Supplementary Table 6**). Top variant rs183091953 was infrequent in HCHS/SOL participants (MAF = 0.002, effective N = 31), and is monomorphic in 1000 Genomes African, European, and South and East Asian reference continental ancestry groups.

## Discussion

In this study, we performed one of the largest GWAS of fatty acids and oxylipins, which contribute to the initiation and resolution of inflammation. Our GWAS identified 19 genetic loci association with 40 oxylipins and fatty acids, including several associations driven by missense or stop gained variants. This work builds on prior GWAS of polyunsaturated fatty acids and a small number of oxylipins and illustrates the shared and distinct genetic architecture affecting fatty acid and oxylipin levels. These results provide insights into the genetic factors influencing circulating fatty acids and oxylipins, and motivate future multi-omics work to better characterize these metabolites and their roles in disease pathways.

Previous studies have reported a wide range in heritability estimates (0% to ∼80%)^12,52,53^ across a variety of metabolites, with significant differences often reported for different metabolite classes.^52^ One previous study of serum oxylipins and polyunsaturated fatty acids in fasting samples from 138 participants from the University College Dublin twin study^54^ reported heritability estimates up to 74% for oxylipins, which is much larger than our oxylipin heritability estimates. However, given that all heritability estimates are population specific, we know that distinct estimates may be driven by genetic and environmental factors, such as diet. For example, the highest heritability estimates in our sample were reported for polyunsaturated fatty acids that are far along the enzymatically regulated biosynthesis pathway from essential dietary fatty acids linoleic acid and alpha linolenic-acid (e.g., arachidonic acid, adrenic acid), whereas the heritability estimates of essential dietary fatty acids linoleic acid and alpha-linolenic acid were much lower.

All of the variants we identified in our main effects GWAS, or high LD proxies, have been previously associated with fatty acid metabolites, phospholipids or other unannotated metabolites, as reported in the GWAS Catalog.^55^ Furthermore, two recent studies in the Framingham and Atherosclerosis in Communities (ARIC) cohorts also identified many of the same loci associated with a variety of oxylipins and fatty acids as we report here.^56,57^ Many of the loci underlying the genetic architecture shared between multiple oxylipins and fatty acids are involved in downstream effects, such as small molecule transportation or signaling, that have important consequences for many endogenous compounds. For example, the *SLCO1B1* locus, which was associated with multiple oxylipins from both n-6 (e.g., arachidonic acid, linoleic acid) and n-3 (e.g., alpha-linolenic acid) fatty acid precursor groups, encodes the hepatocyte-expressed OATP1B1 protein, a membrane influx transporter.^49^ OATP1B1 has both exogenous substrates, including statin drugs, and endogenous substrates like oxylipins, bilirubin, and conjugated steriods.^49,58^ Several variants, including our top variant rs4149056, a missense variant, have been associated with statin-induced myopathy and other related drug interactions^48,49^ due to their impact on OATP1B1 transport activity.^48,59^ Previous studies on the effect of *SLCO1B1* variants on endogenous substate transportation have been limited to bilirubin and were largely inconclusive.^49,60–62^ However, the associations we identified between *SLCO1B1* and many oxylipins motivate the need for future experimental studies to determine whether rs4149056 and other variants also impact oxylipin transportation.

In addition to shared genetic architecture, we also identified 15 loci associated with three or fewer metabolites, indicating that there were also distinct genetic effects influencing circulating fatty acid and oxylipin levels. Among these distinct effect loci, the average percent of trait variance explained by our top variants was 0.94%, suggesting that many of the metabolites associated with these loci may be more polygenic, with additional unidentified small effect variants. These loci are involved in a range of biological functions, including oxylipin synthesis and metabolism (*CYP* genes),^63–65^ and elimination and detoxification of drugs and other endogenous compounds (*UGT2B7*).^66,67^ Indeed, several of these distinct loci may offer insights into the potential identity of putative novel oxylipins. For example, in addition to associations with OATP1B1 transporter protein locus *SLCO1B1* and CYP locus *CYP2C9*, one putatively novel oxylipin was also associated with loci implicated in fatty acid elongation (*PECR*),^68,69^ hydrolysis of activated fatty acids into their non-esterified forms (*ACOT6/ACOT2, HEATR4*),^70^ and catalyzation of the transfer of fucose residues to cell surface acceptor molecules such as glycoproteins, glycolipids, and oligosaccharides (*FUT2*).^71,72^ Collectively, the activities of these associated loci suggest that this oxylipin could be a long chain fatty acid compound or related metabolite, though more formal analyses will be necessary to characterize putative novel oxylipin metabolites. Emerging methods that integrate genomics and metabolomics are showing promise for metabolite annotation,^73^ offering novel means to improve annotation given the ubiquity of large-scale genomics.

Notably, contrary to previous studies,^12,19^ we did not identify associations between genes encoding lipoxygenase (LOX) enzymes (*ALOX5, ALOX12, ALOX12B, ALOXE3, ALOX15, ALOX15B*)^74^ and oxylipins derived from these enzymatic reactions. Our inability to replicate these associations may be due to the composition of oxylipin species quantified in our samples; for example, there were relatively few HETE oxylipins. This finding likely reflects both heterogeneity in the composition of fatty acid and oxylipin profiles in our data due to diet and other factors, as well as limitations in the non-targeted LC-MS methods used to specifically capture and annotate bioactive metabolites. Similarly, while other studies have reported associations between the *ELOVL2* locus and eicosapentaenoic acid, docosapentaenoic acid, docosahexaenoic acid, and adrenic acid,^16,57^ we did not identify those associations here.

However, we did identify two variants (rs28600890-*ADHA1* and rs183091953-*CARS2*) that are monomorphic in at least one other 1000 Genomes continental ancestry reference population, but not in the admixed American HCHS/SOL sample. *ADH1A* top variant rs28600890, which is monomorphic in European and South and East Asian 1000 Genomes reference continental ancestry populations, is in high LD (D’ = 1) with variant rs28864441, which was previously associated with unannotated metabolite levels in an African ancestry population.^75^ Furthermore, by extending our GWAS models to perform two-degree-of-freedom tests to detect both variant and variant-waist circumference-interaction effects on circulating oxylipins, we identified a rare variant specific to admixed American continental ancestry populations near the *CARS2* locus associated with a leukotriene (m/z = 480.25, RT=0.351).

Although this finding requires external replication, particularly considering the low frequency of the top variant, one recent study demonstrated an anti-inflammatory role for *CARS2* in macrophages and smooth muscle cells,^76^ suggesting a biologically plausible link between *CARS2* and the leukotriene oxylipin.

### Strengths and Limitations

Utilizing a high-throughput and sensitive LC-MS method for comprehensively profiling fatty acids and oxylipins and conducted in a sample of over 11,000 participants, this study is one of the largest GWAS of oxylipins and related fatty acids to-date. Moreover, it was conducted in the ancestrally diverse HCHS/SOL cohort, which is important given that most genetic association and metabolomics studies have been historically conducted in European ancestry populations. Indeed, we identified two variants (rs28600890-*ADHA1* and rs183091953-*CARS2*) that are monomorphic in the 1000 Genomes European continental ancestry reference population, but not in admixed American reference populations, thus enabling identification of these signals in the HCHS/SOL cohort.

However, there are also several limitations to this study. First, formal replication of metabolite-genetic locus associations in independent populations is challenging, due to differences in measurement platforms and annotation of metabolites. In particular, we identified multiple significant associations with putative novel oxylipins, and therefore formal replication will not be possible until these can be more definitively annotated. However, the following loci that we identified here were also recently associated with many annotated and novel oxylipin compounds in the Framingham and ARIC cohorts^56,57^: *FADS, SLCO1B1, UGT2B7, PKD2L1, ACOT6, PECR, THEM4, ADH1A*, and multiple *CYP* loci. This supports our results and suggests that the associations between these loci and oxylipins and fatty acids are real signals, though future work will be needed to better characterize the specific oxylipin subgroups that are influenced by each locus. Second, although circulating oxylipin concentrations can become elevated due to secretion or spillover from inflamed tissues,^77^ the actions of oxylipins are likely highly localized and specific to tissue and cell types, which can be challenging to measure in non-experimental settings. Thus, our results may not capture genetic associations that underly tissue-specific oxylipin activity and profiles. However, they still provide insights into systemic oxylipin and fatty acid profiles and the pathways in which they operate.

## Conclusions

Our study revealed shared and distinct genetic architecture underlying correlated and modest-to-moderately heritable fatty acids and bioactive oxylipins. The genetic architecture included loci involved in a variety of biological functions, including fatty acid and oxylipin synthesis, exogenous and endogenous substrate transport and metabolism, and inflammation regulation. These results provide insights into the genetic factors influencing circulating bioactive lipids, and motivate future multi-omics work to better characterize these compounds and elucidate their roles in disease pathways.

## Data Availability

All summary statistics will be available upon publication through the NHGRI-EBI GWAS catalog (https://www.ebi.ac.uk/gwas/).

## Acknowledgements

The authors thank the staff and participants of HCHS/SOL for their important contributions. A complete list of staff and investigators has been provided by Sorlie P., et al. in Ann Epidemiol. 2010 Aug; 20: 642-649 and is also available on the study website http://www.cscc.unc.edu/hchs/.

## Author Contributions

CGD and CLA designed the study. CGD performed the statistical analysis. MJ, TL, and SC generated the oxylipin data. CGD and CLA wrote the manuscript with input from all authors.

